# *In vitro* potency of combinatorial antifungal drugs towards *Candida* isolates from patients

**DOI:** 10.1101/2022.02.28.22271608

**Authors:** Sakshi Agarwal, Smitanjali Parida, Bhuvnesvri Jadoun, Shreya N Patel, Sumit Moolchandani, Himanshu Puniyani, Raviranjan Kumar Pandey, Mandvi Mathur, Sandeep K Shrivastava, Aakanksha Kalra

## Abstract

Candidiasis is a fungal infection (mycosis) caused by an opportunistic yeast with only 8-10 pathogenic species. In spite of the multiple classes of antifungal drugs available to combat these infections, the treatment is hampered by drug toxicity, tolerability, emergence of drug resistant isolates and many more. Combination therapy has been suggested as a possible approach to improve these treatment outcomes. This study is, therefore, aimed at the analysis of the sensitivity of *Candida* isolates obtained from patients towards different classes of drugs affecting multiple pathways in the pathogen life cycle. Four different individual drugs with different mechanisms of action and six different combinations of these drugs at three different proportions (1:1, 1:3, and 3:1) were used for the study. All the drug sensitivity assays were performed by the agar well diffusion method. The results were statistically analyzed using Prism software. Results have shown synergistic effects of drug combinations on the sensitivity of the isolates without much variation in the stoichiometric ratios, thereby providing an alternative to monotherapy to combat the emergence of drug resistant isolates and drying existing drug pipelines.

## 1. Introduction

The last few years have witnessed progressive increase in mycotic infections which are one of the primary causes of morbidity and mortality especially in immunocompromised and severely ill patients. *Candida* is a fungal pathogen which is usually a member of normal flora but transits to pathogenic form by several virulence factors like adherence to host tissues and medical devices, secretion of extracellular hydrolytic enzymes etc. *(Deorukhkar et al., 2014)*. The species is responsible for various clinical manifestations including overgrowth as well as life threatening disseminated infections. *Candida* species have been reported as the fourth most common nosocomial bloodstream pathogen by National Nosocomial Infections Surveillance System (NNISS) with a mortality rate of about 45% *(Deorukhkar et al., 2014)*. Till date, more than 150 species of *Candida* are known with only 10 species responsible for the disease with *C. albicans* being responsible for majority of the infections. However, recent studies have shown the increase in incidences of infection due to non-albicans *Candida* species like *Candida auris, Candida tropicalis, Candida glabrata, Candida parapsilosis, Candida krusei* and *Candida lusitaniae (Abi*□*Said et al., 1997; Da Costa et al., 2009)*. The fatality of fungal infections in immune-compromised conditions has been recently observed with mucormycosis (black fungus) and *Candida* (white fungus) infections (*Soni et al., 2021)*. These infections spread worldwide in both COVID-19 positive patients as well as patients recovered from COVID-19. Studies have shown the presence of oropharyngeal Candidiasis in patients suffering from COVID-19 *(Salehi et al., 2020)* and *C. auris* infections in critically ill COVID-19 patients *(Chowdhary et al., 2020)*.

Several non-albicans species are inherently resistant or acquire resistance to commonly used antifungal drugs. Factors like severe immunosuppression, prematurity, use of broad-spectrum antibiotics, and usage of antimycotic drugs are associated with this change *(Deorukhkar et al., 2014)*. Six different classes of antifungal drugs with different modes of action are available which selectively eliminate fungal pathogens from a host with minimal toxicity to the host. These include Azoles, Allylamines, Polyenes, Echinocandins, Nucleotide Analogue and Penicillin derivatives. Amphotericin and Azoles have been the choice of drugs for patients suffering from mucormycosis and *Candida* infections during the COVID-19 outbreak *(Song et al., 2020)*. However, numerous recent studies have shown the emergence of drug resistant *Candida* strains and isolates. This along with increase in disease causing species has led to a dire need for development of newer drug strategies. One of these novel drug strategies is the combinatorial therapy consisting of multiple drugs affecting different pathways which have proven to be much more efficacious than a single counterpart. It offers numerous advantages such as no or vey less toxicity analysis, lower treatment failure rate, lower case-fatality ratios, fewer side-effects than monotherapy, slower development of resistance, and thus less money needed for the development of new drugs *(Cuenca-Estrella, 2004)*. Thus, in view of this, the present study is focused on determining the drug sensitivity profiles of *Candida* isolates obtained from patients in Rajasthan, India. Further, we have also analyzed the efficacy of drug combinations of multiple drugs in three different stoichiometric ratios towards these isolates. All the results were appropriately statistically analyzed. Our results have clearly shown that all drug combinations are much more efficacious than a single drug towards these isolates without much variation observed at different stoichiometric ratios.

## 2. Materials and Methods

### 2.1 Candida isolates and culture conditions

*Candida* isolates used in the study were kindly provided by MCRD-CIRD (Microbial Culture Repository Division – Centre for Innovation, Research & Department), Dr. B. Lal Clinical Laboratory Pvt. Ltd., Jaipur. These isolates were extracted from various infectious human samples such as urine, blood, pus and vaginal swabs obtained from Candidiasis patients in Rajasthan. Patient’s demographic details such as sex, age and clinical information were recorded and used for analysis. The study consists of a total of 17 isolates which were cultivated and maintained on YEPD medium (Hi Media Laboratories, Mumbai, India) (unless specified) at 30□. The isolates were confirmed macroscopically by visualizing the colony morphology (creamy colony in color) on YEPD agar plates as well as microscopically using Indian ink (Nigrosin) staining.

### 2.2 Preparation of Antifungal drugs and drug combinations

A total of four drugs, Itraconazole, Amphotericin B, Micafungin and Terbinafine, belonging to different classes were used in the study. All the drugs were prepared in DMSO as per CLSI guidelines. Itraconazole and Amphotericin B were used at a concentration of 13.5 mg/ml and 0.33 mg/ml while Micafungin and Terbinafine were used at a concentration of 0.33 mg/ml and 0.435 mg/ml. DMSO was used as a negative control for all the assays.

Depending on the results obtained from drug sensitivity assays using individual drugs, a total of six drug combinations were prepared at three different stoichiometric assays. The drug combinations used for the study are shown in Figure 1.

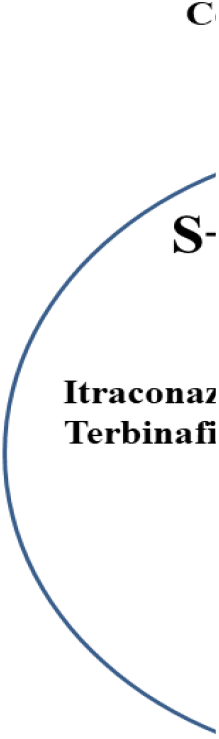

### 2.4 Antifungal drug sensitivity

All the drug sensitivity assays were performed as per standard procedures in accordance with NCCLS guidelines by agar well diffusion assay. Briefly, the isolates were primarily inoculated in Sabouraud Dextrose Broth (SDB) and incubated at 30□ for 24 hrs. These grown cultures were then diluted to an O.D. of 0.2 and then swabbed on SDA plates. Wells were punctured in these plates and 50µL of the prepared drug/drug combination was added into wells. Wells containing DMSO were used as negative control. Plates were incubated for 24 hrs at 30□ and the clear zone obtained around the well, known as the zone of inhibition (ZOI), was measured by zone measuring scale.

### 2.5 Statistical analysis

The mean ZOI ± standard deviation of all isolates for a drug or a drug combination was calculated and was used for the comparison of the significance of drug combinations to individual drugs as well as combinations at various proportions. All the statistical analysis was done by unpaired t test using PRISM version 5.0.

## 3. Results and discussions

### 3.1 Demographic analysis of the isolates

Demographic analysis of the isolates was done on the basis of gender, age group, sample type as well as species causing the infections (Figure 2). The details of the isolates obtained are summarized in the supporting data (Table S1). As it is evident from the results, *Candida* infections are not gender biased and affect both males and females (Figure 2a). However, previous studies have shown that females are slightly more prone to *Candida* infections owing to their higher susceptibility towards UTIs (Urinary Tract Infections) *(Behzadi et al., 2015; Dan et al., 2002)*. The deviation in our results from the previous studies could be attributed to the smaller number of isolates used in the study (*Loster et al., 2016; Strati et al., 2016)*. The analysis of these infections in different age groups suggested almost similar infectivity rate in all age groups with slight increased rates in older age groups owing to weaker immune systems (Figure 2b). This is well in agreement with previous studies suggesting that *Candida* infections mostly occur in immunocompromised patients, so it can occur in any age group who have weaker immune response *(Flevari et al., 2013; Barchiesi et al., 2017; Alrayyes et al., 2019; Laupland et al., 2005)*.

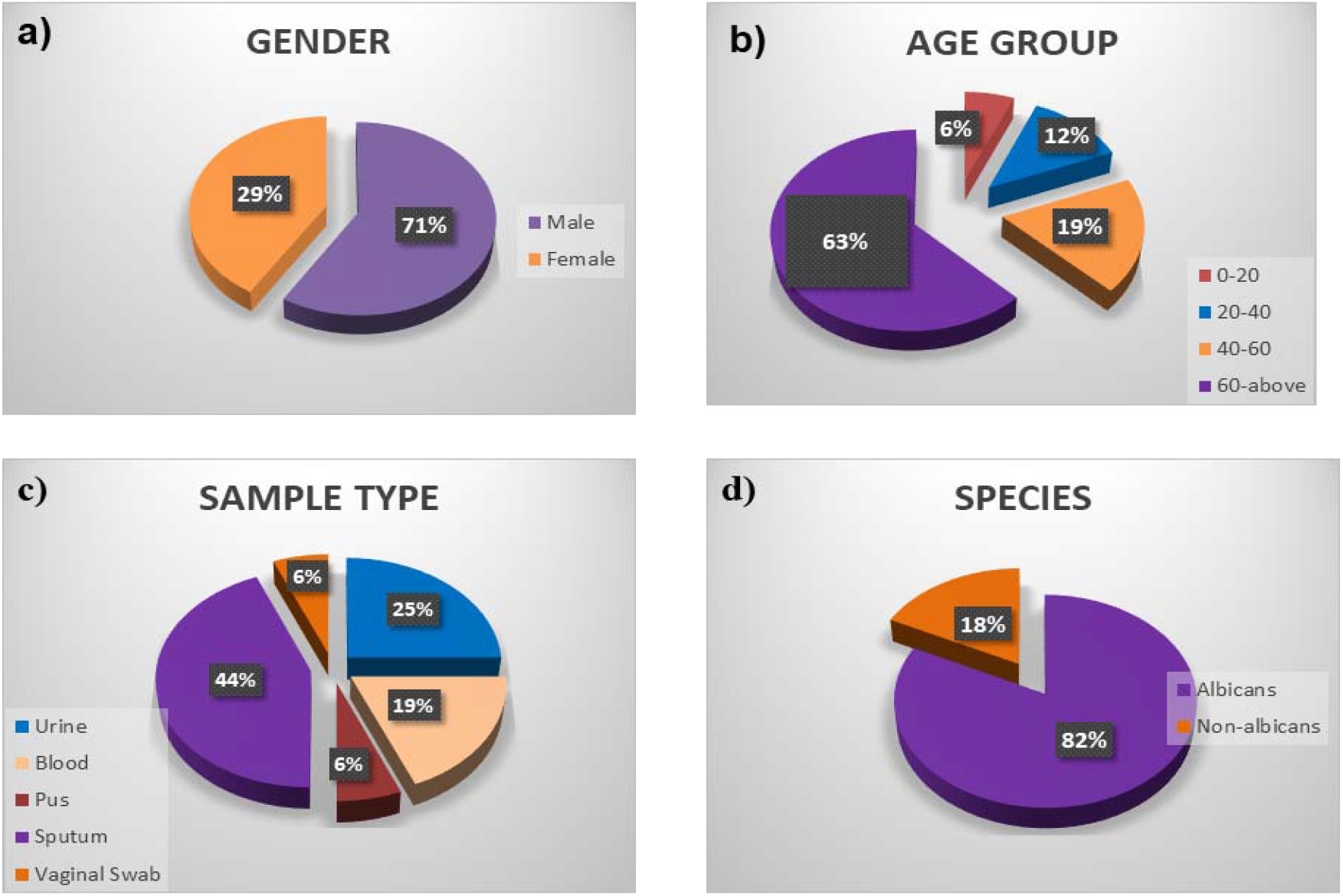

Our results showed that majority of samples were obtained from the patient’s sputum and urine, indicating that *Candida* majorly affects respiratory and urinary systems in the human body (Figure 2c). It can also be correlated from previous studies suggesting that the organism majorly affects lungs causing diseases like pneumonitis *(Pendleton et al., 2017; Huffnagle et al., 2018*). Besides, *Candida* based UTI infections are also very prevalent mostly observed in females as vaginal candidiasis, vulvovaginal candidiasis caused by *Candida albicans (Behzadi et al., 2015; Rodriguez et al., 2020; Tortelli et al., 2020; Achkar et al., 2010)*. Isolates obtained from blood demonstrate the severity of the disease (Figure 2c). Evidences have also suggest the occurrence of *Candida* pneumonia and Candiduria in severely immunocompromised individuals with disseminated disease *(Dermawan et al., 2018; Harris et al., 1999)*. Although the emergence of infections caused by non-albican species is increasing, the majority of the infections in this study were caused by albican species only (Figure 2d). This albican species bias has been observed previously as well from a number of studies (*Kim et al., 2011; Babic et al., 2010; Makeinen et al., 2018)*. However, infections caused by non-albican species such as *auris, glabrata*, etc. are rising continuously which are also causing an increase in emergence of drug resistance *(Bhattacharjee, 2016; Crabtree & Prather, 1933; Glöckner & Cornely, 2015; Deorukhkar et al, 2014; Fu et al., 2017)*.

### 3.2 Drug sensitivity of isolates towards monotherapeutic drugs

Four different antifungal drugs belonging to different classes were used for the study. Drug sensitivity analysis by standard well diffusion assay was done with DMSO as the negative control. Representative images of drug sensitivity assay are shown in the supporting data (Figure S2) and as per CLSI guidelines 17mm ZOI was taken as the cut-off value. On the basis of results shown in Table 1, it is clear that all the isolates were sensitive to Amphotericin B and Micafungin while only 59% and 53% isolates were sensitive to Terbinafine and Itraconazole respectively. The sensitivity of the isolates towards Amphotericin B and Micafungin have also been previously reported to 99.5% and 100% respectively *(Bitew & Abebaw, 2018; Santhanam et al., 2013)*.

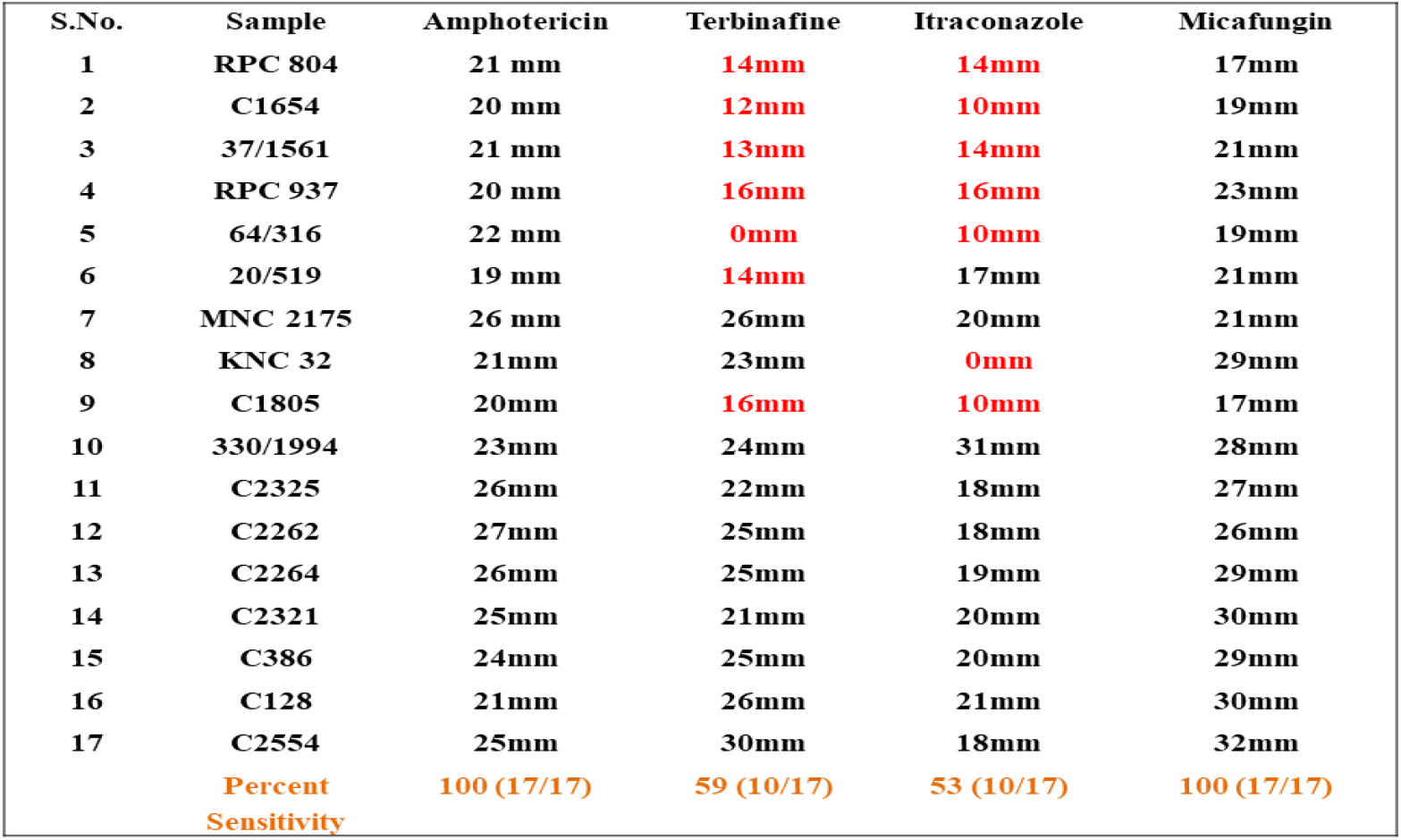

Similar results were observed with patients suffering from oral Candidiasis or autoimmune C*andida* dystrophy *(Rautemaa et al., 2008)*. Similarly, emerging resistance was observed for *Candida* isolates towards Itraconazole *(Santhanam et al., 2013; Rautemaa et al., 2008; Darma et al., 2020)*. Also, emergence of resistant has been observed with isolates towards Terbinafine *(Jensen et al., 2015)*, in patients suffering from Balanoposthitis caused by *Candida albicans (Hu et al., 2016)*. These results along with previous studies clearly indicate the emergence of drug resistant isolates in the area under the study and elsewhere towards Azoles & Terbinafine. Thus, alternate options need to be evaluated to combat the infections caused by the pathogen.

### 3.3 Drug sensitivity of isolates towards combinatorial therapy

Studies so far have indicated that individual drugs may not be the best choice in the current scenario. Multiple studies have illustrated drug combinations to be more efficacious preventing the emergence of resistant strains, lowering the concentration of drugs and treating critically ill patients rapidly *(Ahmed et al., 2014)*. Not just towards *Candida* infections, combinatorial drugs have proven to be effective in other infections as well such as *C*haetomium species (*Sun et al., 2019)*, invasive aspergillosis *(Martin-Pena et al., 2014)*, invasive mold diseases in hematologic patients (*Candoni et al., 2015)*. We, therefore, have analyzed the drug sensitivity of isolates towards drug combinations at three different stoichiometric ratios. The drugs to which more than and less than 60% isolates were sensitive were considered as S and R respectively. Representative images of drug sensitivity assay are shown in the supporting data (Figure S3). The drug combinations were used to evaluate the synergistic effects of drugs in combinations. On the basis of results shown in Table 2, 3 and 4, Itraconazole and Terbinafine were individually not effective to all the isolates but their combination was especially effective when used at 1:1 combination. Similarly, all the other five combinations showed statistically significant synergistic effects towards the growth of the isolates.

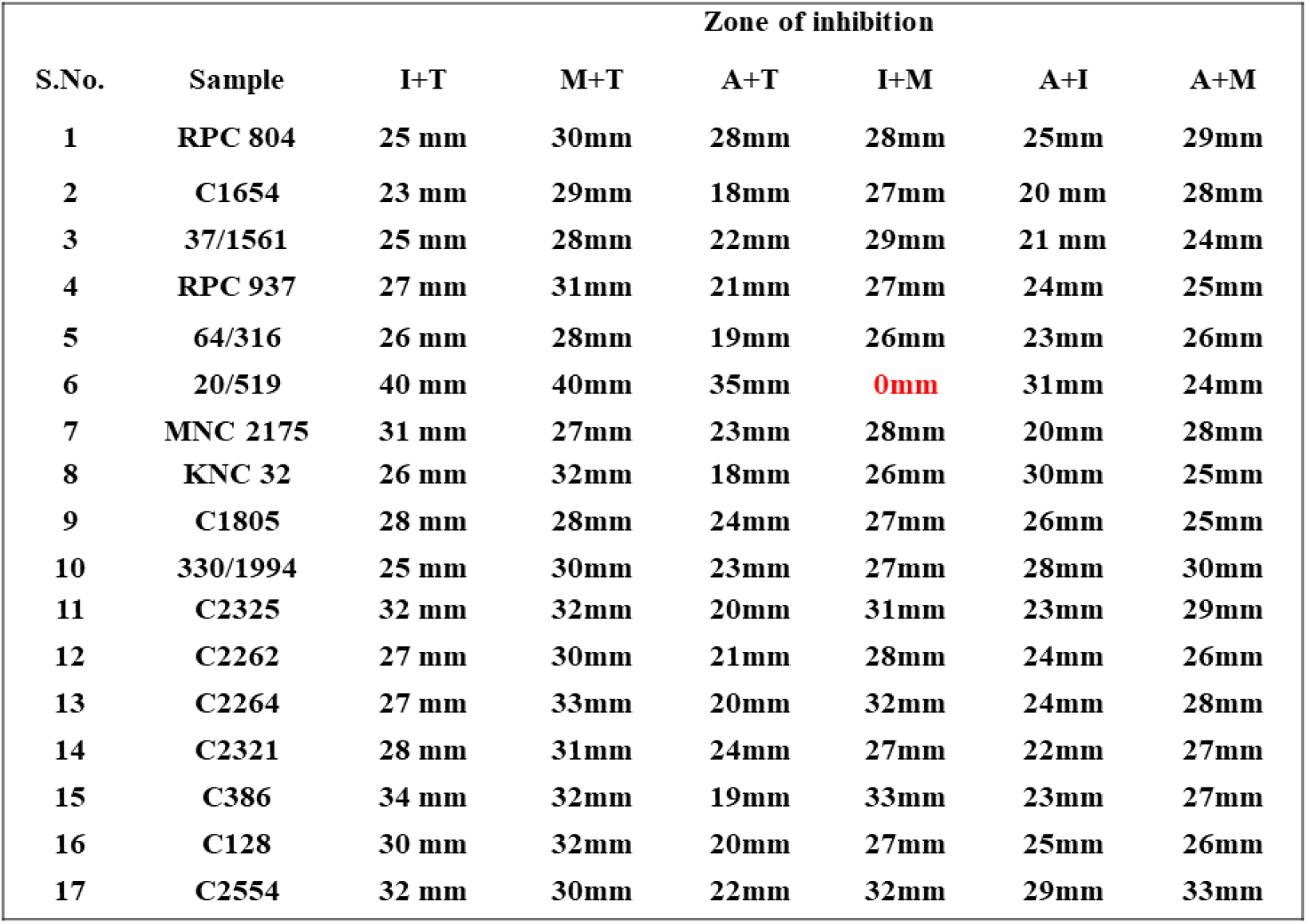

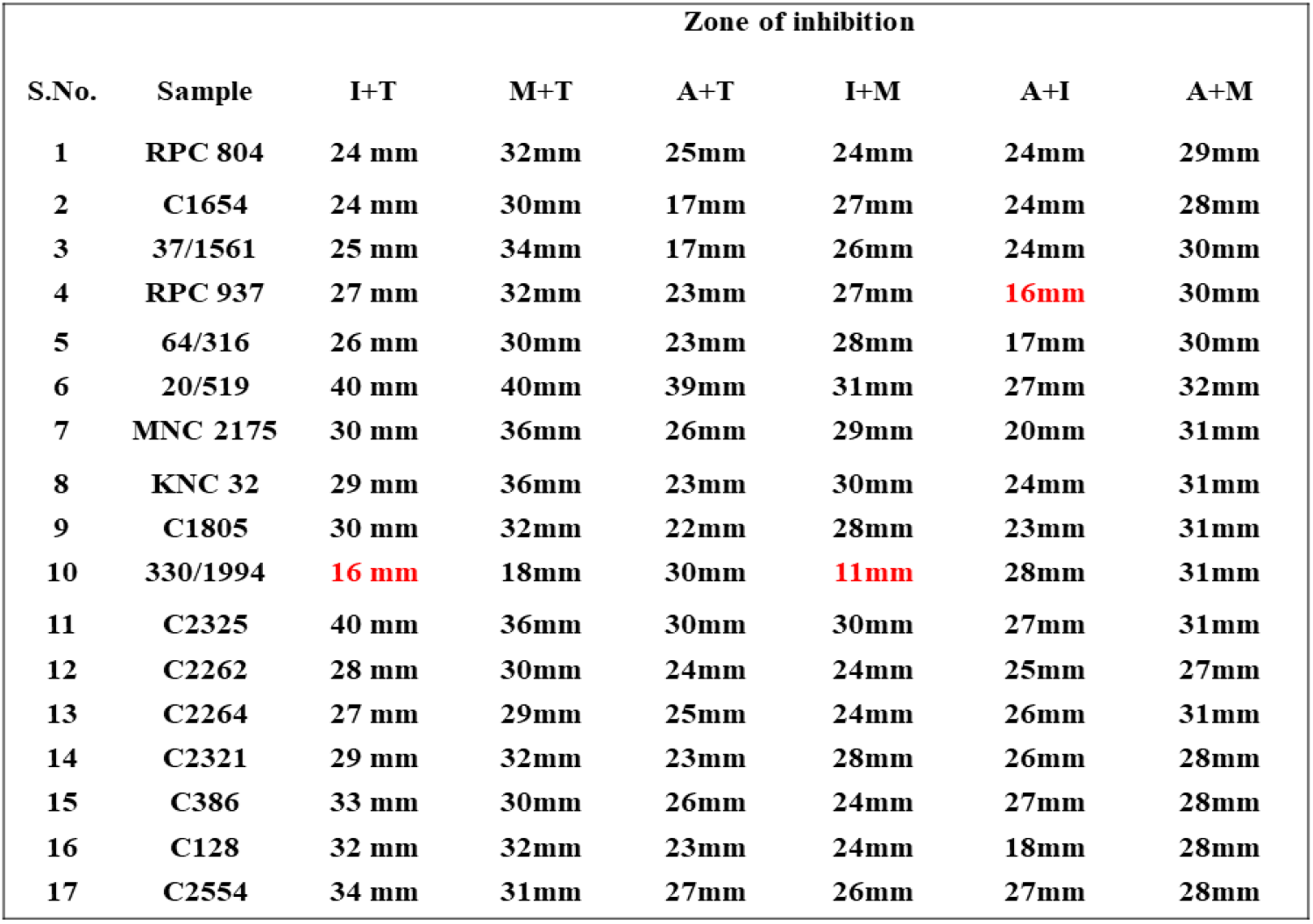

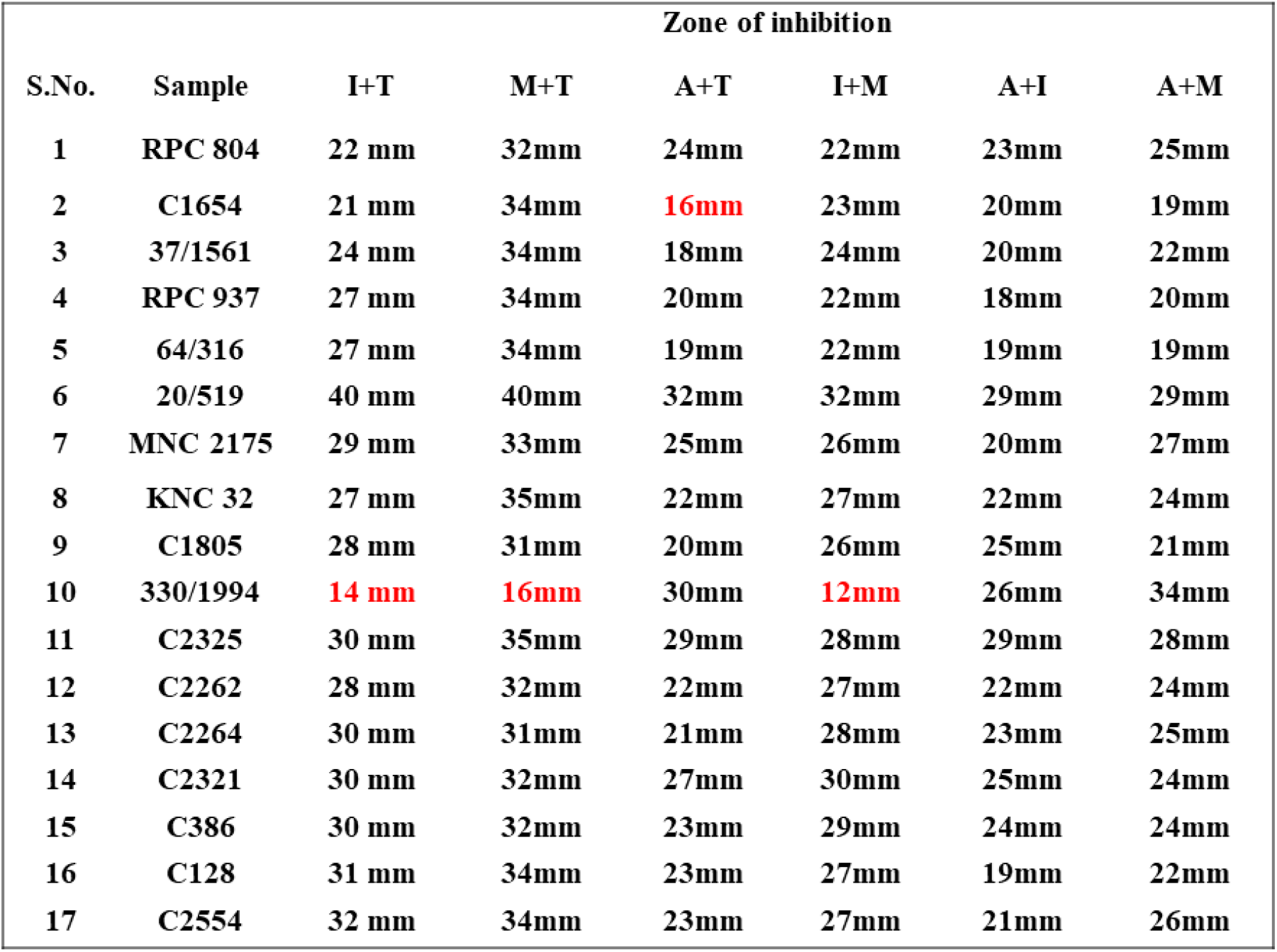

These results are well in agreement with numerous previous studies which have been discussed here. Studies have shown the active and synergistic interaction of azoles with Terbinafine, Amphotericin, and Echinocandins (micafungin) against *Candida* isolates *(Barchiesi et al., 1997; Odds, 1982; Roling et al., 2002; Sugar et al., 1995)*. Another study has reported that a combination of Terbinafine and azole resulted in a fungicidal action against *C. albicans*, although the drugs were fungistatic when they were used individually *(Ghannoum & Elewski, 1999)*. Studies have also shown simultaneous inhibition of different fungal cell targets, giving synergism between Echinocandins (cell wall active) and Amphotericin. Antifungal drug combinations have also been used against planktonic and sessile cells of *Candida albicans (Touil et al., 2018)*. The combinations showed a higher cure rate, fewer relapses, and less nephrotoxicity than monotherapy with Amphotericin *(Johnson et al., 2004)*. Similarly, Terbinafine has also shown synergistic effects due to their mechanism of actions, when used in combination with different drugs like azoles against *Candida albicans & Candida glabrata* infections *(Perea et al., 2002; Hay, 1999)*. Thus, it can be concluded that combination therapy could provide an effective alternative to monotherapy for patients with invasive infections that are difficult to treat due to multi-resistant species.

### 3.4 Statistical analysis of combinatorial drugs at different proportions v/s monotherapy

All the results obtained from drug sensitivity analysis were statistically analyzed as described in materials and methods using Prism version 5.0. The drug combinations were compared to the efficacy by individual drugs along with the three stoichiometric ratios of a combination. Mean ± Std. Dev. of all the combinations and individual drugs are shown in Figure 3. For all the combinations, it was observed that the combinations were much better than monotherapy; however, no significant difference was observed between stoichiometric ratios except in the case of A + T and A + M combinations.

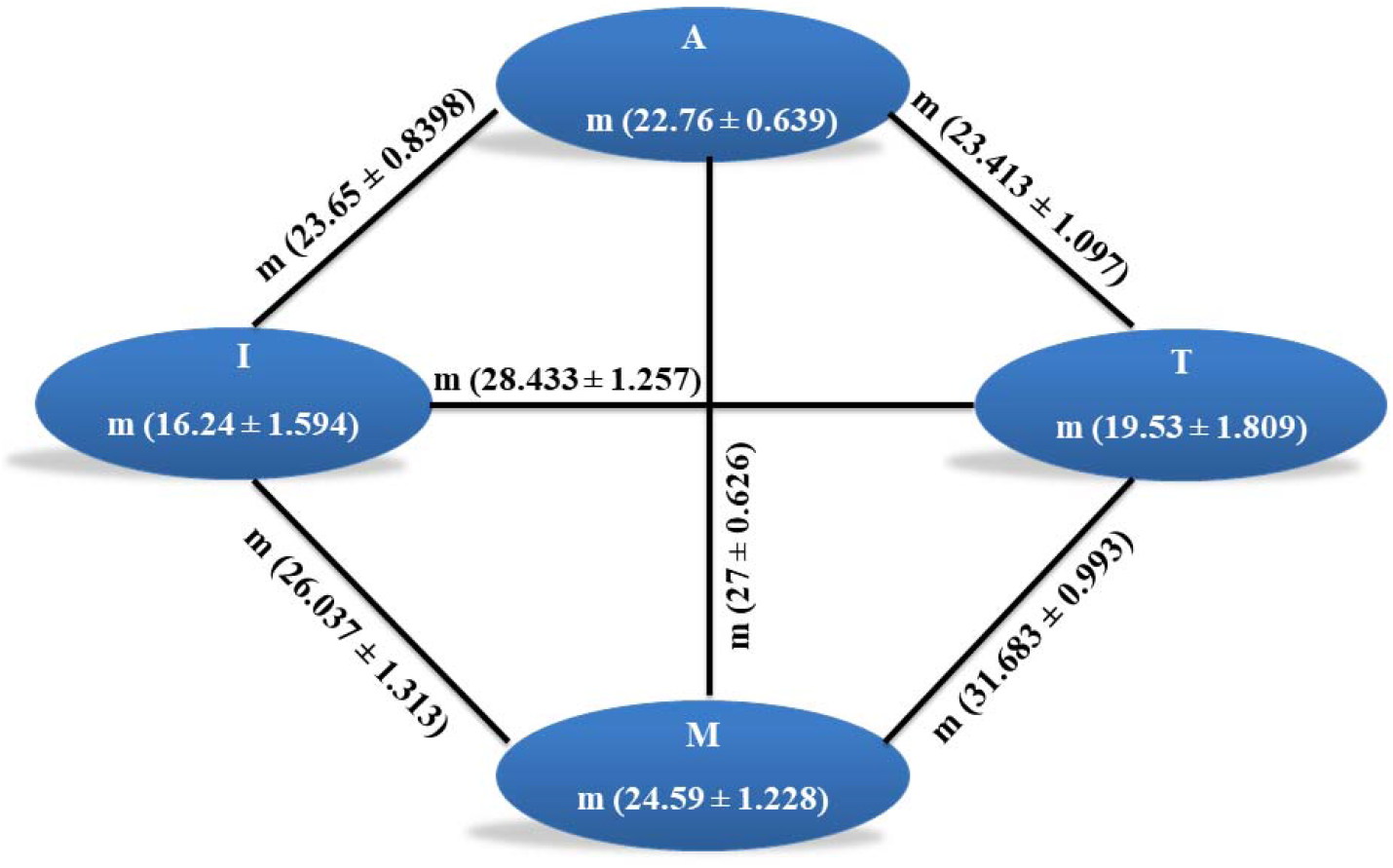

In the A+T combination, the mean was significantly different in 1:3 proportion while insignificant in other proportions as well as to individual drugs. In the A+M combination, the mean for 1:1 and 1:3 was significantly different from the individual drug, and when used in the combination of 3:1, no difference was observed. All the p values obtained for different drug and drug combinations are summarized in Figure 5. A complete analysis of sensitivity of isolates towards different drug combinations is shown in Figure 4.

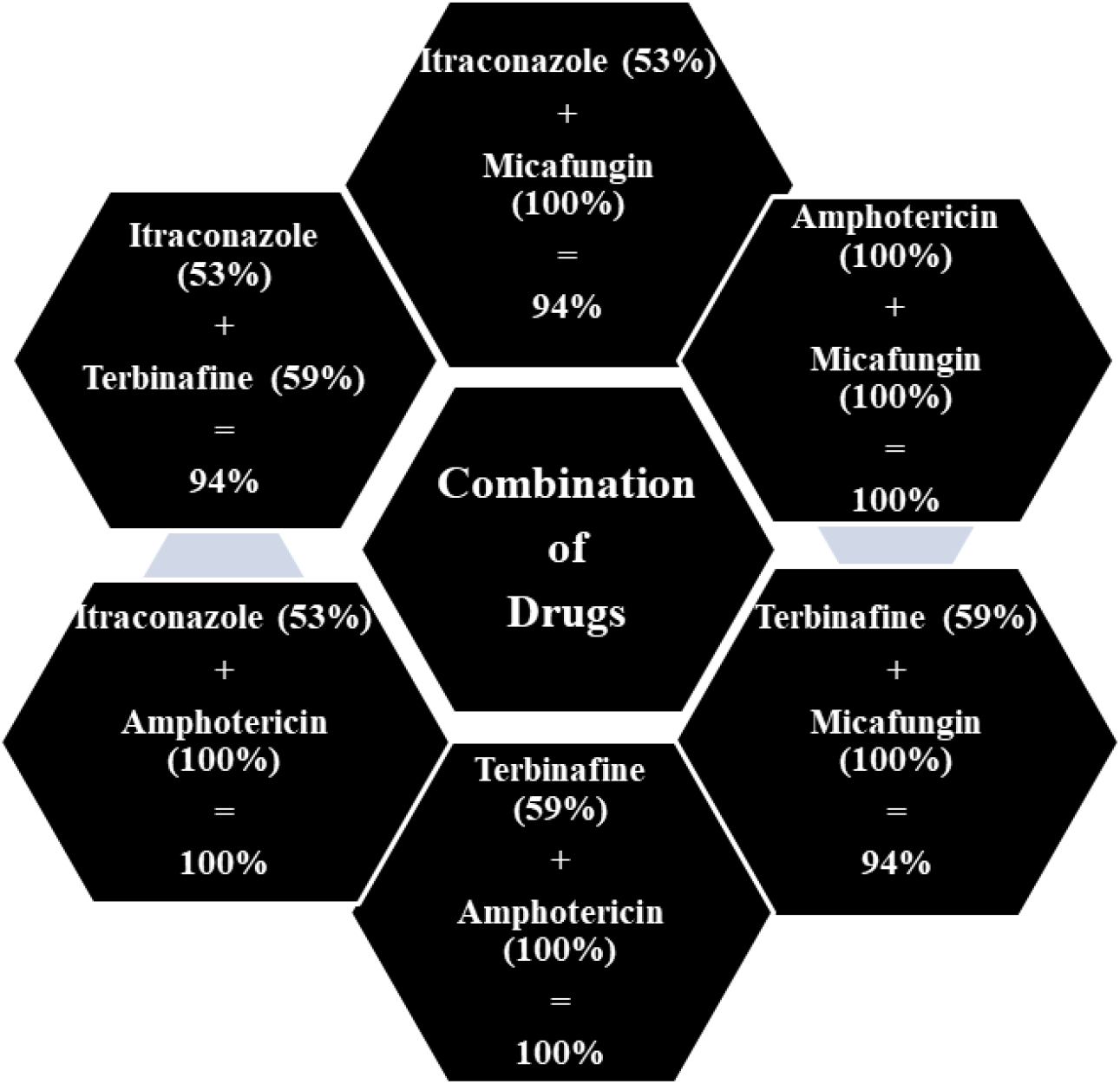

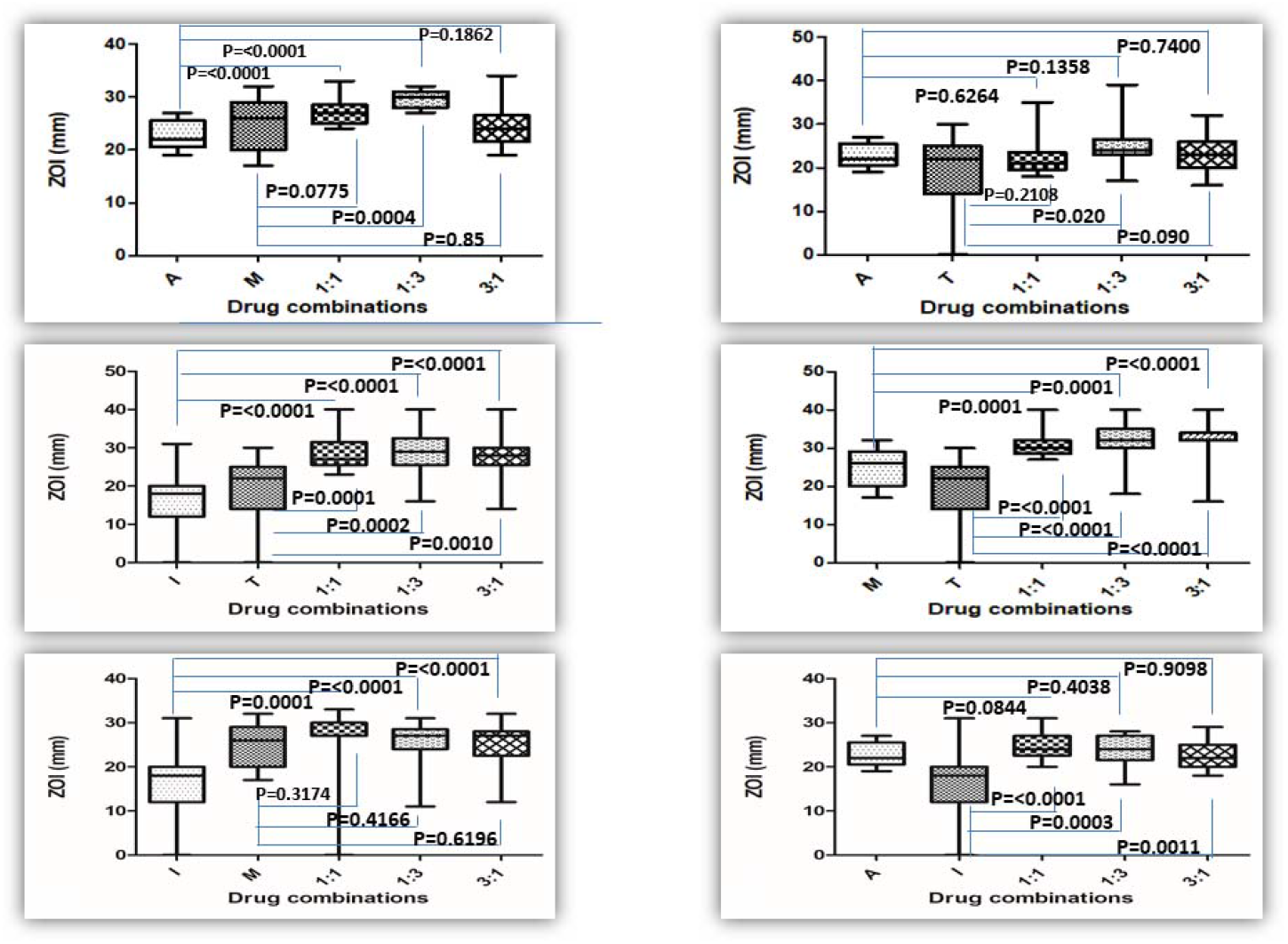

## 4. Conclusion

The study is focused on the drug sensitivity analysis of *Candida* isolates towards various drugs and their combinations. Four different classes of drugs used for the study belonged to different classes with a different mechanism of action (Itraconazole, Amphotericin B, Micafungin and Terbinafine). Results have shown that the monotherapy drugs were not as efficacious. Thus, combinatorial drugs were prepared and analyzed at three different stoichiometric ratios.

The results from this study showed that different isolates showed maximum sensitivities to the combinatorial drugs at all proportions. Most of the combinations were statistically significant from drug monotherapy except for Amphotericin B.

No statistical significance was observed between the three different proportions of drug combinations. Combination therapy could provide an alternative to monotherapy for patients with invasive infections that are difficult to treat due to the emergence of resistance.

## Data Availability

All data produced in the present work are contained in the manuscript

## Acknowledgments

The authors acknowledge the Intramural Research Scientific Committee, Dr. B. Lal Institute of Biotechnology, Jaipur for funding the research.

## Declarations

### Consent to participate

The authors have complete consent for the study.

### Consent for publication

The authors have complete consent for publication.

### Availability of data and material

The isolates were kindly provided by Microbial Culture Repository Division, Centre for Innovation, Research and Development, Dr. B. Lal Clinical Laboratory Pvt. Ltd., Jaipur.

### Code availability

Not applicable.

### Authors’ Contribution

SA performed the experiments and wrote the manuscript; SP, BJ, SNP, SM, HP and RKP performed the experiments; RN, SKS provided the isolates; AK designed the research, planned the experiments, performed formal analysis of the results & wrote the manuscript.

